# Optimizing the patient care technician role: a qualitative study on recruitment, training, and career pathways

**DOI:** 10.64898/2026.03.05.26347687

**Authors:** Nasser Aldosari, Muteb Aljuhani, Ali Albzia, Moath Saleh

## Abstract

**Background:** workforce innovative solutions are warranted to respond to the critical global lack of healthcare professionals and sustain delivery of quality patient care. The Patient Care Technician program was one of the strategies implemented to address this challenge by developing a timely pool of workforce who can take non-complex tasks, alleviating workload on other professionals such as registered nurses. However, since this strategy was recently introduced, its implementation and impact on the delivery of care have not yet been sufficiently investigated.

**Objectives:** This study examines the motivations, experiences, and career aspirations of patient care technician students, alongside program providers’ perceptions and challenges in program delivery.

**Design & Methods:** A qualitative phenomenological study was conducted at three institutions in Western Saudi Arabia, including two tertiary hospitals and a university. Semi-structured interviews were conducted with 27 participants; students, lecturers, preceptors, and management staff. Policy documents were also analyzed, and data were examined using Colaizzi’s seven-step method.

**Findings:** Four key themes emerged: (1) reconciling motivations and influences, (2) training dynamics, (3) career advancement, and (4) navigating acceptance. patient care technician students often felt overqualified for their roles, leading to dissatisfaction and career redirection. The program’s effectiveness was hindered by unclear career pathways and the need for greater cultural sensitivity.

**Conclusions:** Recruiting bachelor’s degree graduates for patient care technician student’s roles may be inefficient, as these positions could be filled by lower-degree holders, potentially reducing costs.

**Implications:** To enhance workforce stability, healthcare policymakers should establish clear career pathways, align job roles with educational qualifications, and adapt the program to local cultural and professional expectations. Addressing these issues can optimize the roles of patient care technician students within the healthcare system and serve as a model for similar workforce strategies globally.

## Introduction

The international healthcare workforce is facing increasing challenges due to a critical shortage of healthcare workers [1]. This shortage, identified by the World Health Organization (WHO) as a significant barrier to achieving universal health coverage, is exacerbated by an aging population and the increasing prevalence of chronic diseases [2]. Recent evidence indicates that the global deficit of healthcare workers could reach 10 million by 2030, significantly affecting the ability of health systems to deliver quality care [3].

In response to the critical shortage of healthcare staff, an alternative staffing model involving unlicensed health practitioners has gained popularity across various countries [4]. However, there is considerable variation in the roles, titles, and regulatory standards for these assistants, leading to confusion and potential implications for patient care [5]. Some of these titles include patient care technician, healthcare assistant, nursing aid and nursing assistant. For consistency, the term Patient Care Technician (PCT) will be used in this paper. Some studies suggest that a higher number of PCTs might be associated with lower quality of care [6, 7].

Saudi Arabia, similar to many other countries, has faced significant challenges in maintaining a sufficient healthcare workforce [8]. To address this issue, the Saudi Ministry of Human Resources and Social Development (HRSD), in partnership with the Saudi Commission for Health Specialties (SCFHS), launched a nationwide PCT program. This program aims to train students to assist registered nurses and provide non-complex direct patient care, e.g. taking vital signs and providing dressing and bathing [9]. It was designed by the Health Academy at the SCFHS and based on the American Red Cross Nurse Assistant Training. Applicants were required to hold a bachelor’s degree in Science e.g. biology, chemistry, or physics with a minimum GPA of 3 out of 5, and the program included 48 weeks of theoretical and hands-on training.

Since the PCT has been recently implemented, questions about the program’s effectiveness and the broader impact on the sustainability of health workforce in Saudi Arabia remain unanswered. This study aims to explore the motivations, experiences, and career aspirations of PCT students, alongside the perceptions of program providers and the challenges faced in delivering the PCT program.

### Literature Review

Across international healthcare systems, patient care technicians (referred to healthcare assistant) roles have expanded to decrease nursing costs and support workplace staffing [10]. However, there is a lack of consensus about the professional title for healthcare assistants and whether this group requires professional regulation. The variety of terms for healthcare assistants has resulted in incongruence around their scope of practice and role within the healthcare team, which may adversely impact patient outcomes. Studies show that there are 37 different terms used for healthcare assistants including Certified Nurse Aid and Certified Nursing Assistant being the most common [5].

In the United States, healthcare assistants, often referred to as Certified Nurse Aides or Certified Nursing Assistants, are generally not professionally regulated, with only 12 jurisdictions having professional regulation programs for these roles [5]. Similarly, in Canada, there is significant variation in the titles and regulation of healthcare assistants across provinces and territories. For instance, terms such as Healthcare Aide, Personal Support Worker, and Continuing Care Assistant are commonly used [5].. This lack of standardization complicates the evaluation of their impact on patient care and workforce dynamics [11].

The role of healthcare assistants has been essential in supplementing the nursing workforce, particularly in response to nurse shortages and increasing healthcare demands [12]. However, evidence suggests that introducing nursing assistant roles does not necessarily improve patient outcomes and may not be cost-effective [11, 13]. For example, the addition of unregulated workers has increased the supervision responsibilities for registered nurses, potentially delaying care and reducing overall efficiency [11]. Moreover, there is concern that an over-reliance on unregulated assistants might undermine the quality of patient care [14].

A significant study demonstrated the potential adverse effects of adding assistants in nursing (AINs) to acute care hospital ward staffing. The study found significant increases in adverse patient outcomes, such as failure to rescue, urinary tract infections (UTIs), and falls with injury, following the introduction of AINs. The only significant decrease observed was in mortality rates. These findings suggest that while AINs can help reduce some workloads, their addition might lead to increased risks in patient care due to insufficient training and the complex nature of patient monitoring and care required in acute settings [7].

The debate over the regulation and professionalization of healthcare assistants has been ongoing. Advocates argue that regulation could help clarify the scope of practice, provide career progression opportunities, and improve the quality of care provided [15]. On the other hand, the lack of professional regulation has led to varied educational preparation and inconsistent support for these roles. This inconsistency in training and scope of practice can lead to variability in the quality of care provided by healthcare assistants [10]. The introduction of PCT programs and the expansion of healthcare assistant roles have been positioned as immediate responses to the critical staffing shortages in Saudi Arabia’s healthcare system. While these measures offer short-term relief, their long-term efficacy remains uncertain. There is a pressing need for robust empirical research to assess the impact of PCTs on patient outcomes, workforce dynamics, and healthcare expenditure [13]. Moreover, the success and sustainability of such initiatives are heavily influenced by the cultural context in which they are implemented. Cultural factors play a pivotal role in shaping the acceptance and integration of PCTs within healthcare settings. Evidence suggests that societal values and professional hierarchies can significantly influence how such roles are perceived and legitimized [16].

Notably, the PCT program in Saudi Arabia requires a bachelor’s degree as a prerequisite for training, an entry criterion that has not been clearly justified by the Saudi Commission for Health Specialties (SCFHS), particularly given the foundational nature of the PCT role. One plausible rationale is that the program serves a dual purpose: addressing nursing workforce shortages while also offering a strategic employment solution for an increasing unemployed science graduates (e.g., biology, chemistry, physics).

In conclusion, addressing the educational and regulatory inconsistencies and considering the cultural contexts in which these roles are implemented are essential steps towards optimizing the contribution of healthcare assistants to the healthcare workforce.

### Significance of the Study

As the PCT initiatives are relatively new in many countries, there is limited evidence to guide policy on the effective use of PCTs. The perceptions of stakeholders regarding the PCT program and role remain unexplored. This study aimed to fill this gap by exploring students’ motivations for becoming PCTs, their experiences during the program and their career aspirations. The insights gained from this study are expected to contribute to the literature on the PCT program and its role, potentially informing strategies to mitigate the global health professional’s shortage.

## Materials and methods

### Study Objectives

- Explore the students’ motivations and incentives to become PCTs.
- Identify the students’ positive and negative experiences while undertaking the program.
- Explore the students’ perceptions of their potential role and their future career plans.
- Identify the program providers’ experiences and perceptions of the PCT program.
- Explore the challenges to successfully delivering the PCT program.

### Study methods

This study employed a qualitative phenomenological approach to understand the lived experiences of key stakeholders in the PCT program. Phenomenology offers a valuable way for researchers to delve into people’s experiences and uncover the deeper meanings within them [17]. In this study, an interpretative phenomenological analysis (IPA) approach was used, which blends psychological, interpretative, and personal elements. IPA has been widely adopted in various fields like education, medicine, and psychology. The main focus with IPA is to attempt to truly understand the experiences, perceptions, and views of participants (PCT stakeholders), which aligns with the aim of the study. In this study, the Consolidated Criteria for Qualitative Research Reporting (COREQ) checklist was used [18].

Data were collected using purposive and theoretical sampling techniques. The study took place at two accredited institutions in Western Saudi Arabia: a tertiary referral hospital and a university.

### Study Population

The study involved various stakeholders in the PCT program, including:

- Current PCT students.
- Lecturers and clinical educators involved in the PCT program.
- Management staff, including leaders of the PCT program and the nursing director.

### Eligibility Criteria Inclusion Criteria

- PCT students currently enrolled in the program.
- Students who voluntarily left the program or PCTs who left their jobs after completing the program.
- Senior nurses, management staff, and nurse educators who interacted with PCTs.
- Leaders of the PCT program.

### Exclusion Criteria

- Students in other unlicensed health programs such as phlebotomy and coding.
- Staff or ward nurses who had no direct contact with PCTs.

### Data Collection

Data were collected through semi-structured interviews with participants. Participant recruitment and data collection were conducted between June 2023 and November 2023. The interviews included open-ended questions about the participants’ experiences and perceptions of the PCT program. Each interview lasted approximately 30 to 60 minutes and was digitally recorded. For participants who preferred not to be recorded, note-taking techniques were used. Each participant was interviewed once, with a possible follow-up interview if needed. Pertinent policy documents were used as an additional resource of data.

### Sampling

27 participants were recruited from two different institutions (university and hospital). The initial sample (8 PCTs) was recruited through the program manager, although the manager was not informed of participants’ final decisions to minimize the chance of coercion. Theoretical sampling, which led to the recruitment of 15 more participants, continued until data were saturated.

### Data Analysis

The data were analyzed following Colaizzi’s seven-step method. It consists of transcribing and reviewing recordings, identifying significant statements, formulating and coding meanings, clustering the codes, verifying findings by repeatedly comparing transcripts and themes, describing the themes and essence using participants’ own words for validity, and affirmation of the findings.

The data were securely stored on KAMC’s data storage system for five years after the project’s completion. Digital recordings and printed transcripts were kept in a locked drawer in a locked office to ensure data security.

### Rigor

The topic guide was loosely structured to allow participants to speak freely, aligning with the recommendation to begin interviews with broad, unrestricted conversations to let the interviewee lead. Transcripts were shared with participants for validation after each interview. During the coding stage, the co-authors (MA and AA) coded independently to ensure consistency, resolving any differences through meetings to reach consensus. Additionally, a methodological consultation was sought from a qualitative expert to rigorously adhere to the principles of the phenomenological approach. These measures were implemented to enhance the quality and dependability of the research findings.

### Ethical Considerations

Ethical approval was obtained from two institutes (university: HAPO-02-K-012-2023-06-1669 and hospital: IRB: 23-1098). Potential participants were provided with a Participant Information Sheet (PIS) explaining the study’s nature, objectives, potential risks, and benefits. Written Informed consent was obtained from all participants. The interviews were conducted in private, and all data were anonymized and stored securely. Participants had the right to withdraw from the study at any time, although their data could not be removed once anonymized and part of the dataset. The study adhered to ethical standards to satisfy the two relevant institutions and ensure the privacy, confidentiality, and safety of participants.

## Results

The data analysis reveals several key themes and subthemes related to the perceptions and experiences of the Patient Care Technician (PCT) program participants. The findings are summarized in Fig 1. Codes elicited eight categories, which were further collapsed into four sub-core categories – reconciling training, dynamic career advancement, and navigating acceptance – and a core category of ‘Acquiescing. Each of the sub-core categories will be discussed first before consideration is given to the core category.

**Fig 1.**
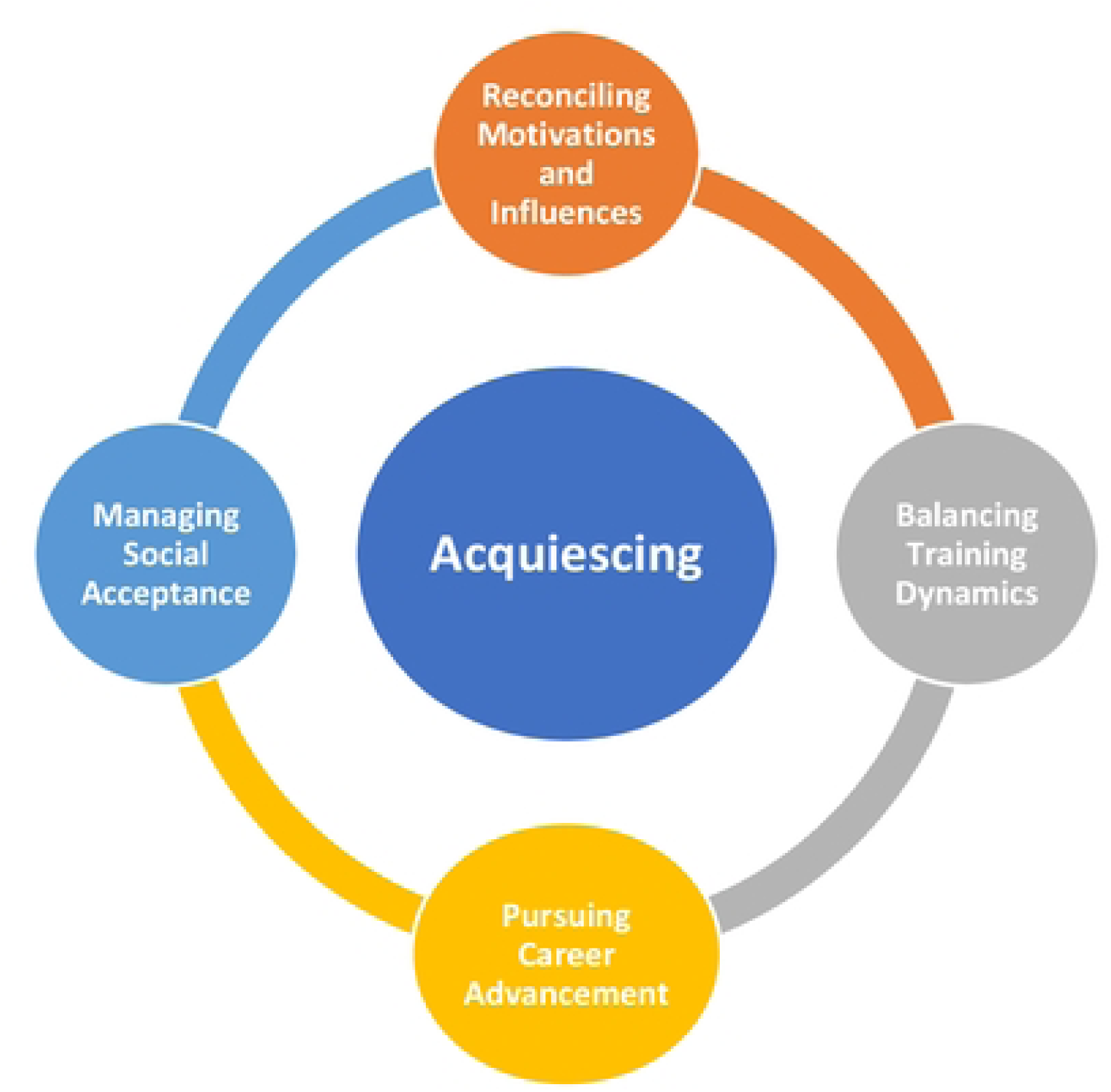
Summary a/ themes identified /ram Interviews.

### Core Category: Acquiescing

The integrative process reflects how participants manage and harmonize various aspects of their experiences in the PCT program to achieve their professional and personal goals.

### Category 1: Reconciling Motivations and Influences

Participants described the multifaceted process of reconciling personal ambitions with external influences when choosing to join the PCT program. Some PCTs had initially aimed for other healthcare professions but turned to the PCT program due to different factors. For instance, one participant mentioned:

> *“It was not the first choice; I applied for the ECG Program but was not accepted. My family influenced me to apply for the PCT program.” - PCT 6*

Social media also have had a twofold effect, providing both encouragement and discouragement. As another participant noted:

> *“Social media influenced me in two directions positively and negatively.” – PCT 3*

This category highlights how participants integrated diverse and sometimes conflicting inspirations and influences into their decision-making process, eventually shaping their professional career choice.

### Category 2: Training Dynamics

The participants’ experiences during the training phase underscored the need to integrate theoretical knowledge with practical skills. Theoretical training was frequently described as intensive and at times overly repetitive. One participant expressed a common sentiment:

> *“The theoretical part was too long compared to the practical part. If it were the other way around with a shorter theoretical period, we would have gained better benefits.” – PCT 16*

Clinical-based training, on the other hand, was highly valued for its hands-on experience. Some participants suggested a more incorporated approach to learning where theoretical and practical training are combined more effectively.

> *“The practical training period was excellent. I wished it could be combined with theoretical training like two days of theory and three days of practical work in hospitals.” – PCT 2*

This theme emphasizes the participants’ aspiration for a cohesive educational experience that better prepares them for their roles.

### Category 3: Career Advancement

A significant theme that emerged was the participants’ strong desire for career progression and further education. Many viewed the PCT role as a stepping stone to more advanced positions within healthcare. One participant (instructor) articulated this aspiration, suggesting:

> *“If there is any chance to improve the program to be a ‘specialist’ it will improve people’s retention in the program.” – PCT instructor*

The lack of clear career pathways was a common concern, with participants expressing frustration over limited opportunities for advancement. Another participant shared:

> *“I will continue as PCT but if there are any opportunities for career development, I will take it.” – PCT 23*

This theme reflects the participants’ efforts to integrate their current roles with their future career aspirations, navigating the healthcare landscape to seek opportunities for growth and development.

### Category 4: Navigating Acceptance

Participants also highlighted the challenges of integrating their professional identity as PCTs within the broader healthcare system and society. Cultural barriers were a significant issue, particularly in performing tasks that are culturally sensitive. One participant noted:

> *“Some competencies are not suitable for female PCTs like perineal care for male patients.” – PCT 8*

Additionally, the lack of recognition and respect for the PCT role within healthcare settings posed a challenge. As one participant remarked:

> *“PCT program and role are not well known in the hospitals. I went to hospitals to ask about the program and they did not recognize the program nor the role.” – PCT 1*

This theme underscores the participants’ ongoing efforts to gain social acceptance and professional recognition, navigating cultural and social landscapes to establish their roles and receive due acknowledgment.

Collectively, the findings underscore the complexity of the PCT experience, revealing a multilayered journey shaped by personal aspirations, systemic limitations, cultural norms, and institutional dynamics. Participants’ motivations to join the program were not merely the result of individual career planning but rather a negotiation between personal goals and external influences, including family expectations and limited academic alternatives.

This nuanced decision-making process challenges simplistic assumptions about career choice in healthcare and calls for a more contextualized understanding of student enrollment patterns.

Training experiences further revealed a critical disconnect between theoretical instruction and clinical practice, with participants advocating for more integrated pedagogical models that reflect real-world demands. Meanwhile, the strong desire for career advancement, despite constrained pathways, reflects both the ambition of PCTs and the systemic failure to provide structured progression routes.

## Discussion

This study is the first to shed light on the perspectives of participants regarding the PCT program in Saudi Arabia. The findings indicate a high level of dissatisfaction among most participants regarding the PCT program. Majority participants felt that the PCT program, in its current form, does not align with their ambitions or academic qualifications. The program primarily targets individuals with bachelor’s degrees in science disciplines who have been unable to find employment in their areas of study, rather than focusing on addressing gaps in the healthcare sector [9].

In contrast, some European countries have adopted a more flexible approach to addressing healthcare workforce shortages. In these countries, there are no strict entry requirements for becoming a healthcare assistant, apart from basic literacy and numeracy skills. The lack of flexibility and inclusivity in Saudi Arabia’s entry requirements for PCT program may limit accessibility and exclude a large pool of potential candidates, thereby undermining the program’s intended purpose. This issue became evident in the early stages of the PCT program, where many potential candidates were rejected for having a bachelor degree other than the targeted ones.

Therefore, to make the PCT program more appealing, it is crucial to broaden the admission criteria in Saudi Arabia. Following the example of other countries, eligibility should be extended to high school and intermediate school graduates to help alleviate workforce shortages in the healthcare sector [12].

The participants also discussed additional factors contributing to their dissatisfaction with the PCT program, particularly those related to motivation, stigma, training dynamics, career advancement, and managing social acceptance.

### Reconciling Motivations and Influences

Participants described a complex interplay of internal ambitions and external pressures, including family expectations, societal norms, and the influence of social media, which shaped their decision towards PCT program. Many participants were driven by a blend of personal aspirations, social influences, and the perceived potential for career advancement. Nevertheless, for some, the PCT program emerged as a secondary option, pursued after encountering barriers to their preferred healthcare career pathways.

Family influence emerged as a significant factor, with some participants feeling pressured to make choices that aligned with their family’s expectations. This pressure may have affected their ability to fully engage with the program, highlighting the tension between personal ambitions and social obligations. These findings highlight the need for a structured career guidance and counselling services to support students to making independent career decisions. These findings are consistent with previous research indicating that healthcare career decisions are frequently shaped by external social and familial pressures, as well as personal goals [19].

Social media has played a multifaceted role in shaping participants’ perceptions of the PCT profession. On one hand, it has provided encouragement and portrayed the PCT profession as an accessible entry point into healthcare. Conversely, it has also reinforced negative stereotypes, leading to a range of mixed perceptions regarding the role. This dual influence aligns with previous evidence indicating that social media can exert both positive and negative effects on public opinion and career-related decision-making in Saudi Arabia [20].

### Stigma

Stigma is a social phenomenon in which individuals are marginalized and devalued by individuals, society, or institutions due to a specific condition, characteristic, or identity [21]. The findings of this study indicate that participants experienced both societal and organizational stigma. Societal stigma was evident in participants’ reports that social media stereotypes showed PCT role as being limited to basic tasks, such as taking vital signs, with no involvement in healthcare decision-making.

Organizational stigma was reflected in the job description of the PCT program in Saudi Arabia, which limit role of health assistants to assisting nurses, requiring that all their tasks be performed under nursing supervision [9]. This is despite the fact that all participants in the program hold an academic qualification equivalent to nursing, namely a bachelor’s degree. In contrast, other programs offered to individuals with a bachelor’s degree in sciences such as the Electrocardiogram Technician program, they have greater professional recognition, autonomy, and participation in healthcare decision-making. This program has a stronger reputation in the community and provide their graduates with more autonomy in their roles compared to those in the PCT program [9].

### Training Dynamics

Participants valued the PCT program’s clinical training but expressed concern about the program’s excessive emphasis on lengthy and repetitive theoretical components (28 weeks theoretical vs 20 weeks hands-on training). They felt this focus left them inadequately prepared for real-world practice. This sentiment aligns with global research highlighting the need for integrated curricula that balance theory with experiential learning to enhance clinical competence [22, 23, 24]. Participants recommended curriculum adjustments to increase hands-on training, simplify theoretical instruction, and improve clinical supervision.

### Career Advancement

Career advancement also emerged as a significant concern, with participants viewing the PCT role as a stepping stone to advanced healthcare careers. However, the lack of structured pathways and specialization opportunities caused frustration, reflecting broader trends in healthcare workforce dissatisfaction [11, 12]. Participants suggested introducing specialized tracks, advanced certifications, and bridging programs to enhance job satisfaction, retention, and the long-term sustainability of the PCT program [25]. These findings highlight the urgent need for curriculum reforms and policy initiatives to strengthen both training quality and career development opportunities.

### Managing Social Acceptance

Participants reported significant challenges in gaining recognition and respect within both the healthcare system and broader society. This was largely due to limited public understanding of the PCT role and cultural sensitivities surrounding gender-specific care tasks. Female participants, in particular, expressed discomfort when performing intimate care for male patients, which reflects broader cultural norms in Saudi Arabia.

These issues align with previous findings from highlighting the influence of cultural barriers and the lack of professional regulation and standardization for PCT roles. This lack of regulation contributes to confusion and inconsistent recognition within healthcare settings. Participants stressed the need for public awareness campaigns, the development of professional practice guidelines, and the implementation of structured orientation programs for both healthcare staff and patients [5, 17]. These initiatives would enhance understanding, acceptance, and the professional integration of PCTs into healthcare environments.

## Conclusion

There is a fundamental disconnect between the program’s structure and the expectations, qualifications, and experiences of its trainees. Rather than serving as a coherent response to healthcare workforce shortages, the PCT program appears to function as a stopgap for graduate unemployment, resulting in dissatisfaction, stigma, and role ambiguity. Issues such as rigid entry criteria, imbalanced training models, limited career progression, and lack of social and institutional recognition reflect deeper systemic and cultural misalignments. These insights challenge the assumption that workforce interventions alone can resolve structural gaps without a comprehensive and context-sensitive design.

### Implications

This study reveals key implications for healthcare workforce policy in Saudi Arabia. The mismatch between program design and participants’ qualifications reflects a policy-practice gap that limits the program’s effectiveness. Cultural and gender norms significantly influence the acceptance and integration of PCTs, suggesting that workforce planning must account for social context. The lack of professional recognition and regulatory clarity weakens the role’s legitimacy within healthcare settings. Additionally, the imbalance between theoretical and practical training highlights the need for more practice-oriented, competency-based education models.

### Recommendations

To improve the PCT program’s relevance and impact, admission criteria should be broadened to include high school and diploma graduates. The curriculum must be restructured to reduce excessive theory and increase hands-on clinical training. Establishing clear career pathways, such as bridging and specialization tracks, is essential to support retention and professional growth. Public awareness campaigns and staff orientation should be used to improve recognition of the PCT role. Cultural and gender sensitivities must be addressed in training and deployment, and formal job descriptions and regulatory frameworks should be implemented to standardize the role and reduce stigma.

### Limitation

Although the study sample included multiple stakeholder groups from two different locations, some limitations should be highlighted when interpreting the findings. The data were generated from a purposive sample of participants within a specific region in Saudi Arabia, which may limit the transferability of the findings to other settings. While efforts were made to capture a diverse range of views, the study reflects the particular experiences and social contexts of those who chose to participate. Additionally, the reliance on self-reported narratives introduces the possibility of recall bias or social desirability bias, especially when discussing sensitive issues such as stigma or cultural norms. Finally, this study primarily foregrounds the voices of PCT trainees and instructors; the absence of perspectives from policymakers, employers, or patients presents a partial view of the program’s broader sociocultural and institutional dynamics.

## Data Availability

The data underlying the results presented in this study are not publicly available due to ethical restrictions and the sensitive nature of qualitative interview data, which may contain information that could compromise participant confidentiality. Data are available from the King Abdullah Medical City Institutional Review Board (IRB) upon reasonable request for researchers who meet the criteria for access to confidential data. Requests may be directed to the IRB Office at King Abdullah Medical City, Makkah, Saudi Arabia.

## Acknowledgments

The authors would like to thank all participants who took part in this study for sharing their experiences and perspectives. The authors also acknowledge the support of the participating institutions for facilitating data collection and providing access to relevant policy documents. Special appreciation is extended to the program coordinators and clinical educators who assisted in coordinating the recruitment process and supporting the research activities.

## Notes

### Competing Interest Statement

The authors have declared no competing interest.

### Funding Statement

The author(s) received no specific funding for this work.

### Author Declarations

Ethical approval for this study was obtained from two institutions in Saudi Arabia. Approval was granted by the university ethics committee (HAPO-02-K-012-2023-06-1669) and the hospital Institutional Review Board at King Abdullah Medical City (IRB: 23-1098). All participants received a Participant Information Sheet and provided informed consent prior to participation. Confidentiality and anonymity were maintained throughout data collection and analysis.

